# Prevalence and factors associated with incompletion of measles and rubella vaccine routine immunization among children aged 18-59 months in Kilimanjaro region, Tanzania

**DOI:** 10.1101/2024.07.02.24309864

**Authors:** David M. Gichogo, Abdul A. Mbezi, Sasi K. Sasi, Vailet Magogo, Edwin J. Shewiyo, William Nkenguye, Florian Tinuga, Sia E. Msuya

**Affiliations:** Institute of Public Health, Department of Community Health, Kilimanjaro Christian Medical University College (KCMUCo), P.O. Box 2240 Moshi, Tanzania; Kilimanjaro Clinical Research Institute (KCRI), Box 2236, Moshi, Tanzania; Institute of Public Health, Department of Epidemiology and Biostatistics, Kilimanjaro Christian Medical University College, P.O. Box 2240 Moshi, Tanzania; Immunization and Vaccine Development Program, Ministry of Health, Dodoma, Tanzania; Department of Community Health, Kilimanjaro Christian Medical Centre (KCMC), P.O. Box 3010, Moshi, Tanzania

**Keywords:** Measles, Rubella, MR vaccine, Vaccination, Children, Prevalence, Tanzania

## Abstract

**Background:** In Tanzania Measles and Rubella (MR) vaccination coverage of 90% in 90% of the regions was achieved in 2018. However, there are councils lagging in MR1 coverage, 38 (19%) councils out of 195 had <90% coverage by the end of 2019 while MR2 coverage is struggling with 99 (51%) of councils having a coverage of <90% at the end of 2019. Kilimanjaro region is among the regions with some councils having MR coverage of <80%. There is a need for information on why a significant proportion of eligible children are not receiving the second dose of MR vaccine.

**Objective:** To determine prevalence and factors associated with incompletion of MR vaccination among children aged 18-59 months in two districts of Kilimanjaro region.

**Methodology:** This study was a cross-sectional study conducted in Moshi urban and rural councils, two out of the seven councils in Kilimanjaro region. The study population was children aged 18-59 months whose parents/ caregivers have been the residents of the respective districts for at least 3 past years. Interviews using questionnaires were used for data collection. Data was entered and analyzed using SPSS version 20.

**Results:** A total of 415 children aged 18-59 months were enrolled. The proportion of children with incomplete MR2 vaccination was 14.2%. Only 33% of the 415 caregivers knew MR vaccine is given at 9 and 18 months and 24% reported unavailability of vaccine at their scheduled visit. Children from Moshi rural council (AOR=2.53, 95% CI =1.36-4.73) had higher odds of MR incompletion. Lower odds of MR vaccine incompletion were among caregivers who were aware on the time for MR vaccination; 9 and 18 months (AOR=0.27, 95% CI=0.10-0.73), caregivers who had knowledge that MR vaccine protects measles, and those who had health promotion talk on the vaccination day.

**Conclusion:** Nearly one in ten (14.2%) children do not complete the recommended two doses of MR vaccines. Strategies to improve awareness and knowledge on timing, frequency and advantage of MR and other vaccines is needed among parents/ caregivers in this setting. Further, improvement in ordering and supply of vaccines at health facilities needs to be improved.

## BACKGROUND

Measles and rubella, both viral infections, are of significant public health concern due to their potential for widespread transmission and severe health consequences, particularly among vulnerable populations such as young children and pregnant women. Globally, measles remains a leading cause of vaccine-preventable deaths among children, with over 140,000 deaths reported in 2018 alone(1). Sub-Saharan Africa, in particular, bears a disproportionate share of the measles burden, with an estimated 155,000 measles-related deaths occurring in the region in 2018 alone(1).

The introduction of measles-rubella (MR) vaccines has played a crucial role in reducing the burden of these diseases. Through widespread vaccination efforts, substantial declines in measles-related deaths have been observed globally. For instance, between 2000 and 2018, measles vaccination prevented an estimated 23.2 million deaths worldwide, with most of the deaths averted in the African region(2). However, despite these successes, outbreaks of measles persist, particularly in regions with inadequate vaccination coverage.

Tanzania, like many other countries in sub-Saharan Africa, has reported measles outbreaks, despite efforts to control the disease through vaccination. In Tanzania, MR vaccination is administered to children under five years in two doses, with the first dose given at 9 months and the second dose at 18 months, according to the Tanzanian routine vaccination schedule(3). Tanzania has observed an increase in number of measle cases from 2019, with about 12,253 cases reported between 2019 – 2022(4). Children aged 1-4 years had the highest proportion of laboratory confirmed cases compared to other groups. Michael et al (2024) also showed that, MR unvaccinated children had 2.6 higher odds of getting measle outbreak compared to fully vaccinated children. Currently Tanzania is number 9 leading country with measles outbreak(5).

Suboptimal measles vaccination coverage is the main cause of the reported outbreaks by increasing the pool of susceptible children over time. The 2019 data from the Integrated Vaccination Data (IVD) program have shown that despite the availability of the vaccine, there is an incomplete uptake of MR vaccination, with the second dose facing particular challenges. Tanzania MR coverage of 90% in 90% of the regions was achieved in 2018 according to DHIS-2 data. By the end of 2019, coverage of MR1 was high (> 95%), but 19% (38 out of 195) of councils had coverage of < 90% (6). There are many councils lagging when it comes to the second dose of MR (MR2), with 51% (99 out of 195) of councils having a coverage of <90% at the end of 2019 representing a significant gap in vaccination coverage(4). Studies from Dar es Salaam and Mtwara have shown 17% and 55% of children under five had not received the second dose of MR vaccine(7,8).

In Asia studies have shown vaccine availability, negative perception on vaccines and poor knowledge on vaccination schedule to be factors associated with incompletion of MR vaccine(9,10). Literature from East Africa reported both health system and caregivers’ characters to influence completion of MR vaccination. Awareness of MR vaccine among caregivers, vaccine hesitancy driven by misinformation or cultural beliefs, myths, negative attitudes, and poverty contribute to the incompletion of the MR vaccination schedule(11). Logistical barriers, such as limited access to vaccination services in remote or rural areas, and distance to facilities also influenced completion of MR vaccine(4,12).

Against this backdrop, this study aims to investigate the prevalence and factors associated with the incompletion of MR vaccine routine immunization among children aged 18-59 months in Kilimanjaro region, Tanzania. The region is among those with MR2 vaccine coverage of < 90% in several councils. By identifying barriers to vaccination uptake and elucidating the context-specific challenges faced by communities, this research seeks to inform evidence-based strategies for enhancing vaccination coverage and advancing progress towards Sustainable Development Goal 3.8, which focuses on achieving universal health coverage, including access to essential vaccines, for all individuals by 2030.

### Specific Objectives

1. To determine the prevalence of MR2 vaccine incompletion among children aged 18-59 months.
2. To identify the factors associated with the incompletion of MR2 vaccination.
3. To assess caregivers’ awareness and knowledge regarding MR vaccination.

## METHODS

### Study Design

This was a community based cross-sectional study conducted from June to August 2022.

### Setting

The study was conducted in two out of seven councils of Kilimanjaro region, namely Moshi municipal council and Moshi district council. The region is situated in northern Tanzania with approximate population of 1,861,934 and 224,780 children U5. Moshi municipal council has a total population of 221,733 people and 25,280 children U5, while Moshi DC has a population of 535,803 people and 61,762 children aged U5 years (Census, 2022). The two districts were selected because they had low coverage where MR2 immunization coverage of <80% according to the Immunization Coverage update for performance of routine immunization of January to August 2021 in Tanzania. Main economic activities done in the areas are tourism, agriculture and manufacturing activities.

### Study Population

The study population was children aged 18-59 months. Parents especially mothers and/ or caretaker of the child-aged 18-59 months provided the information. The study included children whose parents or caretakers were the residents of the two selected councils. Residency in this study was defined as having an address in the selected council and having lived in the area for the past three years prior to the study.

### Sample size and sampling

Multi-staged sampling technique was used to access the study participants. From the list of 53 wards, 21 wards in Moshi municipal and 32 wards from Moshi DC respectively, a sample of 6 wards 3 wards from each district with their respective population of children under 5 years were randomly selected. Random sampling was employed for each selected ward. The sample size for each selected ward was obtained.

From each selected ward, all streets within the ward were listed and 7 streets were drawn randomly from Moshi municipal and 2 villages from Moshi DC from the selected wards. The households were randomly selected and from the households, and a caregiver or parent was interviewed. In situations where there was more than one child with 18-59 months of age all were interviewed.

### Data Collection

Face-to-face interviews with a questionnaire was used at household level to collect information from parents and caretakers of the children aged 18-59 months. The questionnaire was uploaded into Kobo toolbox, an online data kit. Prior to the interview, informed consent was sought from the study participants then the interview proceeded among the consented caregivers. Interviews lasted between 15 and 20 minutes depending on the respondents’ ability to respond to posed questions.

### Study variables

#### Dependent variable

Dependent variable of this study was incompletion of MR2 vaccination according to the immunization card and by report speech.

#### Independent variables

The independent variables of this study included; caretakers’ individual factors (such as fear of adverse events following vaccination, awareness on the need for MR second dose, attending antenatal and postnatal care), health system factors (such as absenteeism of health worker at the health facility, vaccine availability, existence of outreach services, opening days, time of vaccination session, long waiting time), caretaker’ socio-demographic factors (such as occupational status, education level, economic status, marital status and family size), child factors (such as age, sex, child birth rank, orphan hood) and community factors (such as travelling distance to health facility, means of transport and geographical accessibility to health facility).

### Data Analysis

The collected questionnaires from Kobo toolbox were checked manually for its completeness, and after reviewing them the data was transferred and analyzed using SPSS version 20. Data was cleaned by running the frequency of each variable. Categorical variables were summarized into frequency and percent tables. Numeric variables were summarized using mean/ median and their respective measures of dispersion. Bivariate and multivariable analyses were conducted to identify independent factors associated with the incompletion of MR2 vaccine. Crudes Odds Ratios (COR) and Adjusted Odds Ratios (AOR) and 95% Confidence Intervals (CI) were calculated. Variables with p-value of < 10% in bivariate analysis were entered into multivariable logistic regression models.

## RESULTS

A total of 425 children aged 18-59 months were approached. A total of 415 (97.6%) children were enrolled, 10 children were not included because the caregivers were not around at the time of interview.

### Background characteristics of the participants

The median age of the 415 children was 35 months, (IQR 24-46) and 52% were males. Most of the informants were mothers (86.0%) with a median age of 28 years (IQR 25-34). About half (50.4%) of the informants had secondary education or higher and most of them were married (65.5%). The common occupational activity of informants was informal employment (47.7%) where the majority earned less than 50,000 Tanzanian shillings per month (56.6%), **(Table 1)**.

**Table 1:** Social demographic characteristics of children aged 18-59 months and caregivers in Moshi urban and rural districts, Kilimanjaro region (N=415)

| Variable | Frequency | Percentage |
| --- | --- | --- |
| **District of enrollment** |  |  |
| Moshi urban | 226 | 54.5 |
| Moshi rural | 189 | 45.5 |
| **Age of the child(months)** |  |  |
| Median (IQR) | 35(24-46) |  |
| 18-24 | 107 | 25.8 |
| 25-36 | 128 | 30.8 |
| 37-48 | 104 | 25.1 |
| 49-59 | 76 | 18.3 |
| **Sex of the child** |  |  |
| Male | 214 | 51.6 |
| Female | 201 | 48.4 |
| **Informant** |  |  |
| Mother | 357 | 86.0 |
| Father | 11 | 2.7 |
| Other relatives | 47 | 11.3 |
| **Informant's age** |  |  |
| Median (IQR) | 28(25-34) |  |
| 18-24 | 86 | 20.7 |
| 25-34 | 232 | 55.9 |
| 35-44 | 50 | 12.1 |
| 45+ | 47 | 11.3 |
| **Marital status of the informant** |  |  |
| Not married | 143 | 34.5 |
| Married | 272 | 65.5 |
| **Educational level of the informant** |  |  |
| Primary level and below | 206 | 49.6 |
| Secondary level and above | 209 | 50.4 |
| **Occupation of the informant** |  |  |
| Non employed | 143 | 34.5 |
| Informally employed | 198 | 47.7 |
| Formally employed | 43 | 10.4 |
| Others | 31 | 7.1 |
| **Average income of the informant (TZS)** |  |  |
| ≤50000 | 235 | 56.6 |
| >50000 | 180 | 43.4 |
| **Number of people in the household total** |  |  |
| ≤5 | 330 | 79.5 |
| >5 | 85 | 20.5 |
| **Number of children in the household** |  |  |
| ≤4 | 401 | 96.6 |
| >4 | 14 | 3.4 |
| **Distance to health facility** |  |  |
| <2km | 340 | 81.9 |
| >2km | 75 | 18.1 |
| **Obstacles to reach the health facility** |  |  |
| Yes | 38 | 9.2 |
| No | 377 | 90.8 |

### Health facility characteristics and Caregivers’ awareness and knowledge on MR vaccination

Majority (82%) of the 415 parents/ caregivers reported to live less than 2 kilometers to a health facility that provides vaccination services. About 1.9% of 415 parents/caregivers of the enrolled children reported that they did not find the health worker at the vaccination post on the scheduled date of vaccination and about quarter (24.1%) reported they attended for vaccination, but the vaccines were out of stock, (**Table 2**).

**Table 2:** Background characteristics of health facilities and awareness, knowledge of caregivers on MR vaccination in Moshi urban and rural districts, Kilimanjaro region (N=415)

| Variable | Frequency | Percentage |
| --- | --- | --- |
| **Awareness of MR vaccine** |  |  |
| Yes | 367 | 88.4 |
| No | 48 | 11.6 |
| **Knowledge on time of MR vaccination (n=367)** |  |  |
| 9 months | 31 | 8.4 |
| 9 and 18 months | 120 | 32.7 |
| I don't know | 216 | 58.9 |
| **Awareness of importance of MR2 vaccine (n=367)** |  |  |
| Yes | 139 | 37.9 |
| No | 228 | 62.1 |
| **Measles as a vaccine protected disease** |  |  |
| Yes | 366 | 88.2 |
| No | 49 | 11.8 |
| **Vaccination status by card (n=216)** |  |  |
| **MR2** |  |  |
| Yes | 195 | 90.3 |
| No | 21 | 9.7 |
| **Vaccination status by recall (did not have card, n=199)** |  |  |
| **MR2** |  |  |
| Yes | 161 | 80.9 |
| No | 38 | 19.1 |
| **MR2 status by card and recall (N=415)** |  |  |
| Yes | 356 | 85.8 |
| No | 59 | 14.2 |
| **Absenteeism of health worker on vaccination day** |  |  |
| No | 407 | 98.1 |
| Yes | 8 | 1.9 |
| **Unavailability of the vaccine on vaccination day** |  |  |
| Yes | 100 | 24.1 |
| No | 315 | 75.9 |
| **Promotion talk on vaccines when attending for PNC** |  |  |
| Yes | 168 | 40.5 |
| No | 247 | 59.5 |

PNC: Postnatal care

Nearly one in ten (11.6%) of the 415 parents/ caregivers had no awareness of the MR vaccine. Of 367 who were aware, many of the caregivers (58.9%) did not know the time of MR vaccination, and only 33% knew MR vaccine is given twice, at 9 and 18 months as per vaccination schedule in Tanzania (Table 2). Few (37.9%) were aware of the importance of MR2 vaccine, (**Table 2**).

### Prevalence of MR vaccine incompletion

A total of 59 children out of 415 (14.2%) had not completed MR2 at the time of the study. Out of 415 children enrolled in the study, caregivers of 216 children had vaccination cards, 21 children (9.7%) were not vaccinated with MR2. **(Table 2)**.

### Factors associated with MR2 incompletion

Children enrolled in Moshi district council were three times more likely to be unvaccinated for MR2 vaccine compared to children enrolled in Moshi urban (COR=3.20, 95% CI=1.77-5.79). Likewise male children were more likely to be unvaccinated compared to female children (COR=2.02 95% CI=1.13-3.59), (**Table 3**).

**Table 3:**
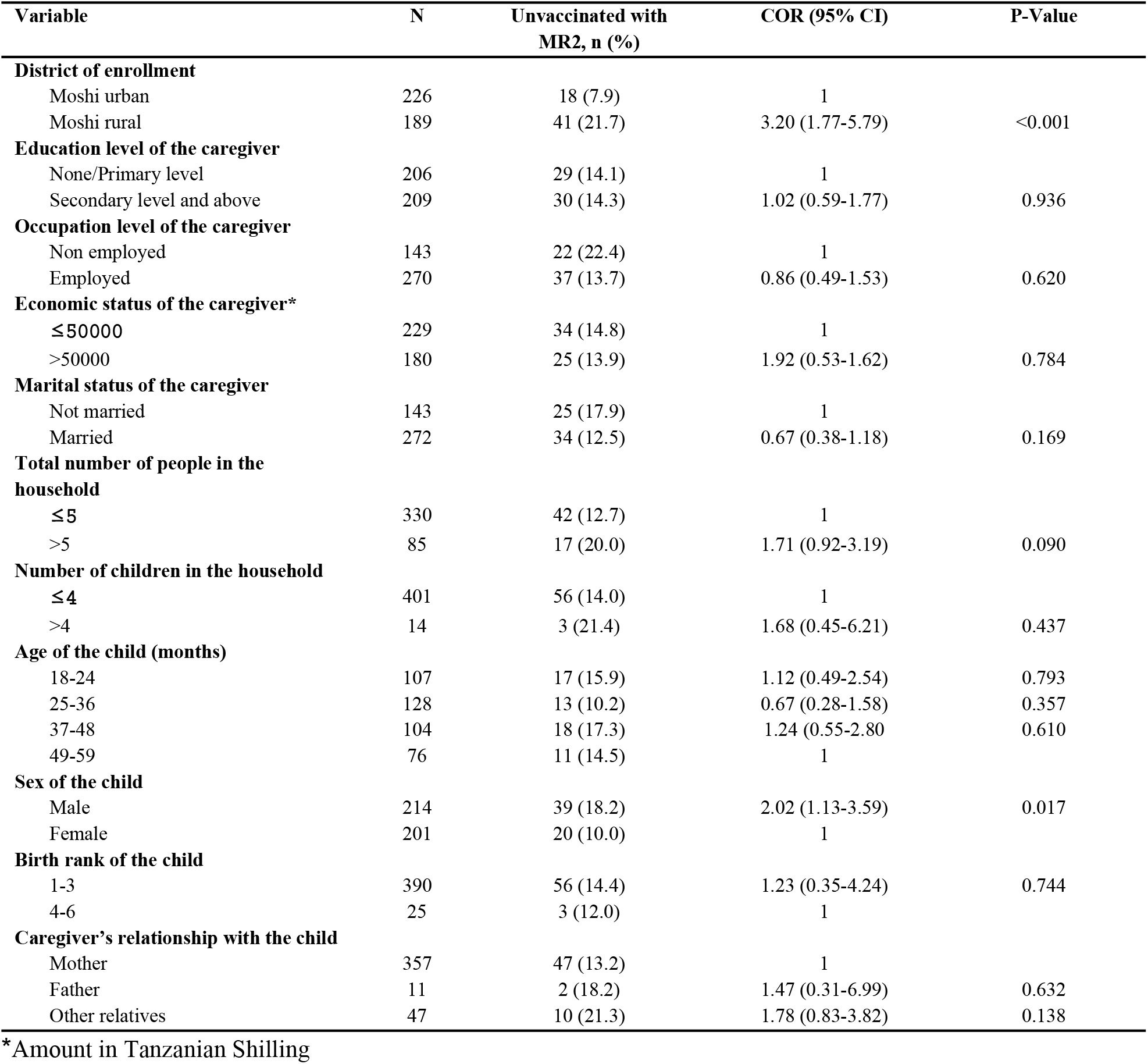
Bivariate analysis of socio-demographic characteristics associated with incompletion of MR2 vaccine in Kilimanjaro region.

Caregivers who did not receive counseling or health promotion talks postnatal care had less odds of completing MR2 vaccination than others. Caregivers who were not aware on importance and schedule of MR vaccination had 4 times higher odds of having a child with incomplete MR vaccination than others (COR=4.38, 95% CI=1.91-10.05), **(Table 4)**.

**Table 4:** Bivariate analysis of the health facility, caregivers’ and community factors associated with the incompletion of MR2 vaccine.

| Variable | N | Unvaccinated with MR2, n (%) | COR (95% CI) | P-Value |
| --- | --- | --- | --- | --- |
| **Awareness of MR vaccine** |  |  |  |  |
| Yes | 367 | 50 (13.6) | 0.68 (0.31-1.50) | 0.341 |
| No | 48 | 9 (18.8) | 1 |  |
| **Knowledge on time of MR vaccination (n=367)** |  |  |  |  |
| 9 months | 31 | 13 (41.9) | 1 |  |
| 9 and 18 months | 120 | 13 (10.8) | 0.17 (0.07-0.42) | <0.001 |
| I don't know | 216 | 24 (11.1) | 0.17 (0.08-0.40) | <0.001 |
| **Awareness of importance of MR2 vaccine (n=367)** |  |  |  |  |
| Yes | 139 | 7 (5.0) | 1 |  |
| No | 228 | 43 (18.8) | 4.38 (1.91-10.05) | <0.001 |
| **Measles as a vaccine protected disease** |  |  |  |  |
| Yes | 366 | 47 (12.8) | 1 |  |
| No | 49 | 12 (24.5) | 2.20 (1.07-4.52) | 0.032 |
| **Distance to nearest health facility** |  |  |  |  |
| <2km | 340 | 43 (12.6) | 1 |  |
| >2km | 75 | 16 (21.3) | 1.87 (0.99-3.55) | 0.054 |
| **Obstacles to reach the nearest health facility** |  |  |  |  |
| Yes | 38 | 7 (18.4) | 1 |  |
| No | 377 | 52 (13.8) | 0.71 (0.30-1.69) | 0.438 |
| **Absenteeism of health worker on vaccination day** |  |  |  |  |
| No | 407 | 57 (14.0) | 1 |  |
| Yes | 8 | 2 (25.0) | 2.05 (0.40-10.39) | 0.388 |
| **Unavailability of the vaccine on vaccination day** |  |  |  |  |
| Yes | 100 | 20 (20.0) | 1.77 (0.98-3.20) | 0.060 |
| No | 315 | 39 (12.4) | 1 |  |
| **Promotion talk on vaccine when attending for PNC** |  |  |  |  |
| Yes | 168 | 15 (8.9) | 1 |  |
| No | 247 | 44 (17.8) | 2.21 (1.18-4.41) | 0.012 |

### Multivariable analysis of the factors associated with the incompletion of MR2 vaccine

In multivariable analysis, children enrolled in Moshi rural were two times more likely to be unvaccinated with MR2 vaccine compared to those enrolled in Moshi urban (AOR=2.53, 95% CI =1.36-4.73). Likewise, awareness on the time for MR vaccination, children with caregivers who were aware that ages for MR vaccination are 9 and 18 months and those who were not aware were less likely to be unvaccinated with MR2 vaccine compared to those who were aware of only 9 months as the age for MR vaccination (AOR=0.27, 95% CI=0.10-0.73), (AOR=0.17, 95% CI=0.07-0.44) respectively **(Table 5)**.

**Table 5:** Multivariable logistic regression on factors associated with the incompletion of MR2 vaccine.

| Variable | Adjusted OR (95% CI) | P-Value |
| --- | --- | --- |
| **District of enrollment** |  |  |
| Moshi municipal | 1 |  |
| Moshi DC | 2.53 (1.36- 4.73) | 0.004 |
| **Sex of the child** |  |  |
| Male | 1 |  |
| Female | 0.51 (0.27-0.97) | 0.040 |
| **Measles as a vaccine protected disease** |  |  |
| Yes | 1 |  |
| No | 1.56 (0.61-3.96) | 0.350 |
| **Knowledge on time of MR vaccination (n=415)** |  |  |
| 9 months | 1 |  |
| 9 and 18 months | 0.27 (0.10-0.73) | 0.010 |
| I don't know | 0.17(0.07-0.44) | 0.001 |
| Others | 0.25(0.09-0.73) | 0.010 |
| **Awareness of the importance of MR2 vaccine (n=367)** |  |  |
| Yes | 1 |  |
| No | 2.89 (1.17-7.16) | 0.022 |
| **Promotion talk on vaccine when attending PNC** |  |  |
| Yes |  |  |
| No | 1.24 (0.61-2.51) | 0.541 |

## Discussion

Our study found that 14.2% of children had not completed the MR2 vaccination, with an overall vaccination coverage of 85.8%. This rate is higher than the national average of 74% and exceeds the coverage reported in community studies conducted in southern rural Tanzania (44.2%) and urban Tanzania (83%)(8,13). The higher MR2 coverage observed in our study can be attributed to multiple immunization campaigns conducted across various districts in the Kilimanjaro region, which have enhanced access to vaccination services for children in the community.

However, significant disparities in vaccination coverage exist between rural and urban areas. Children in rural areas were found to have twice the odds of missing MR2 vaccination compared to their urban counterparts. This disparity is critical, considering that the majority of the children in the Kilimanjaro region reside in rural areas, thus highlighting significant gaps in immunization coverage. This finding is consistent with studies from other low-resource settings, where geographic and socio-economic barriers often impede access to vaccination services(14,15).

Our study also identified caregiver awareness and knowledge as significant factors influencing MR2 vaccination coverage. Caregivers who were aware of the importance of MR2 vaccination and knew the correct vaccination schedule were significantly less likely to have children who missed the MR2 dose. This aligns with previous research in Tanzania and other settings, which underscores the importance of parental education and awareness in improving vaccination uptake(16,17).The high level of unawareness observed in our study, with 62.1% of caregivers unaware of the importance of the MR2 vaccine and 58.9% unaware of the vaccination schedule, underscores the need for enhanced health education initiatives.

The role of postnatal care (PNC) visits in improving vaccination coverage is evident in our findings. However, about 10% of caregivers reported not receiving counseling or health promotion on vaccination during their PNC visits. This represents a missed opportunity to educate caregivers on the importance of complete immunization. Studies have shown that integrating immunization education into routine PNC services can significantly improve vaccine uptake(18,19).

In terms of health system factors, the availability of vaccines and healthcare worker presence at vaccination posts were crucial. Approximately 24.1% of caregivers reported vaccine stockouts during scheduled visits, which is a significant barrier to vaccination completion. This issue is echoed in other studies from low- and middle-income countries (LMICs), where logistical challenges in vaccine supply chains hinder immunization efforts(20,21).

Furthermore, our study’s findings suggest that community-level interventions and improved healthcare infrastructure are necessary to address these gaps. Ensuring consistent vaccine availability and enhancing health worker training and motivation are essential steps toward achieving higher vaccination coverage. Additionally, targeted outreach programs in rural areas can help bridge the coverage gap between urban and rural populations.

The findings from our study align with the broader literature on vaccine coverage and highlight the multifaceted nature of immunization challenges. Studies have shown that comprehensive approaches, combining health education, improved healthcare delivery, and robust supply chain management, are essential for enhancing vaccination rates(22,23).

For example, a study conducted in Nigeria found that improved training for health workers and enhanced community outreach programs significantly increased vaccination rates in rural areas(14). Similarly, research in Ethiopia indicated that integrating health education programs into routine health services improved caregiver knowledge and vaccine uptake(24). In India, addressing logistical barriers by ensuring consistent vaccine supply and reducing travel distances to health facilities was shown to improve vaccination coverage(25).

In summary, our study highlights the significant gaps in MR2 vaccination coverage in the Kilimanjaro region, with a particular focus on rural areas. Enhancing caregiver awareness through health education, ensuring vaccine availability, and addressing logistical barriers are crucial for improving MR2 vaccination rates. These findings underscore the need for multifaceted interventions to achieve higher vaccination coverage and better health outcomes for children in Tanzania.

## CONCLUSION

The study aimed at assessing the prevalence and factors associated with incompletion of MR2 vaccine among children aged 18-59 months whereas a proportion of (14.2%) of children had not received MR2 dose. Knowledge on MR vaccine, awareness on the importance of MR2 dose, awareness of measles as a disease protected by vaccination and health promotion talks significantly contributed to the incompleteness of MR2 dose. Intervention measures thus should aim at improving community education at all levels to reduce the burden of unvaccinated children.

## RECOMMENDATION

With regard to improving MR2 coverage in Kilimanjaro region, the following recommendations should be considered so as to attain the recommended coverage of >95% by 2025.

- More health promotion talks should be offered regarding postnatal vaccinations whereas caregivers should be educated on the importance of vaccines and the proper schedule for administration of all vaccines for their children. Also, these talks should be extended to the community level so as to reach out even those caregivers who find no importance of vaccinations.
- Health care workers should consider screening vaccination status of children who visit health facilities for other purposes out of vaccination and consider vaccinating those who have missed as per Tanzania immunization guideline.
- The government together with the Ministry of Health could also make efforts to make vaccination knowledge more available in as many different platforms as possible like mass media, social media, religious centers and many more so as to reach a lot of people with this knowledge.

## Data Availability

All relevant data are within the manuscript and its Supporting Information files

## Ethics statement

The ethical clearance was obtained from the Ethical Committee of the Kilimanjaro Christian Medical University College Research Ethics and Review Committee (CRERC) in Moshi **(UG 53/2022)**. Permission was obtained from the President’s Office, Regional Administration, and Local Government (POLARG), from the regional administrative secretary and regional medical officer office. Purpose of the study, benefit, right to refuse participation or leaving the study was explained to each participant before the interview. The study participants were informed that the data will be kept confidential by using codes and not personal identifiers such as names and is meant only for the purpose of study. Verbal consent was regarded as sufficient to be used in the study.

## Patient Consent for Publication

Not Applicable

## Acknowledgement

We are grateful to all participants who participated in one way of this work and the data collection team

